# Sex differences in ADHD diagnosis and clinical care: A national study of population healthcare records in Wales

**DOI:** 10.1101/2023.10.20.23297324

**Authors:** Joanna Martin, Kate Langley, Miriam Cooper, Olivier Y. Rouquette, Ann John, Kapil Sayal, Tamsin Ford, Anita Thapar

**Affiliations:** Centre for Neuropsychiatric Genetics and Genomics, Cardiff University, Cardiff, UK; Wolfson Centre for Young People’s Mental Health, Cardiff University, Cardiff, UK; School of Psychology, Cardiff University, Cardiff, UK; Neurodevelopmental Service, Cwm Taf Morgannwg University Health Board, Merthyr Tydfil, UK; Institute of Suicide Prevention and Mental Health, Swansea University, Swansea, UK; Centre for ADHD and Neurodevelopmental Disorders Across the Lifespan, Institute of Mental Health, Nottingham, UK; Unit of Mental Health and Clinical Neurosciences, School of Medicine, University of Nottingham, UK; Department of Psychiatry, University of Cambridge, Cambridge, UK

**Keywords:** ADHD, sex differences, neurodevelopmental conditions, mental health

## Abstract

**Background:** Population-based studies have observed sex biases in the diagnosis and treatment of attention deficit hyperactivity disorder (ADHD). Females are less likely to be diagnosed or prescribed ADHD medication. This study uses national healthcare records, to investigate sex differences in diagnosis and clinical care in young people with ADHD, particularly regarding recognition and treatment of other mental health conditions.

**Method:** The cohort included individuals diagnosed with ADHD, born between 1989 and 2013 and living in Wales between 2000 and 2019. Routine primary and secondary healthcare record data were used to derive diagnoses of ADHD and other neurodevelopmental and mental health conditions, as well as ADHD and antidepressant medications. Demographic variables included ethnicity, socioeconomic deprivation, and contact with social services.

**Results:** There were 16,458 individuals diagnosed with ADHD (20.3% females, ages 3–30 years), with a male-to-female ratio of 3.9:1. Higher ratios (4.8:1) were seen in individuals diagnosed younger (<12 years), with the lowest ratio (1.9:1) in those diagnosed as adults (>18). Males were younger at first recorded ADHD diagnosis (mean=10.9 vs 12.6 years), more likely to be prescribed ADHD medication, and younger at diagnosis of co-occurring neurodevelopmental conditions. In contrast, females were more likely to receive a diagnosis of anxiety, depression, or another mental health condition and to be prescribed antidepressant medications, prior to ADHD diagnosis. These sex differences were largely stable across demographic groups.

**Conclusion:** This study adds to the evidence base that females with ADHD are experiencing later recognition and treatment of ADHD. The results indicate that this may be partly because of diagnostic over-shadowing from other mental health conditions, such as anxiety and depression, or initial misdiagnosis. Further research and dissemination of findings to the public are needed to improve awareness, timely diagnosis, and treatment of ADHD in females.

---

Attention deficit hyperactivity disorder (ADHD) is a common and impairing neurodevelopmental condition. In childhood, it is more frequently observed in males than females (Willcutt, 2012), and this sex bias is especially marked in referred clinical samples, compared to screened population-based samples (Faraone et al., 2015). Studies of adults report less of a male bias, with many community or general population studies, reporting a 1:1 sex ratio (Faheem et al., 2022). Registry-based studies of children in Denmark and Sweden have found that ADHD tends to be recognised and diagnosed on average at a later age in females (Dalsgaard et al., 2019; Martin et al., 2018; Wimberley et al., 2020). This observed later age at diagnosis is consistent with the less prominent male bias seen in adults. In this paper, we use the terms male and female, as epidemiological and observational studies typically rely on information about sex assigned at birth, rather than gender identity.

Although ADHD prevalence is similar across countries, its recognition, diagnosis and treatment varies across countries, time, raters, and study methodologies (Polanczyk et al., 2007; Rydell et al., 2018; Willcutt, 2012). There is evidence that ADHD is under-diagnosed in the UK (Young et al., 2021), as well as growing recognition that ADHD may be especially under-diagnosed in females (Quinn & Madhoo, 2014; Young et al., 2020). Even when a diagnosis is made in females, treatment with ADHD medication is often further delayed or not provided for younger females, with somewhat less pronounced sex differences in prescriptions for adults with ADHD (Dalsgaard et al., 2014; Kok et al., 2020; Russell et al., 2019).

Further studies are needed to more deeply examine sex biases in current diagnostic practices and clinical care of ADHD. Diagnostic overshadowing from various mental health conditions may delay recognition of ADHD in females (Quinn & Madhoo, 2014; Young et al., 2020). However, to date there is little empirical research that has examined this important possibility.

A variety of demographic factors could also influence the likelihood of diagnosis and treatment, acting in combination with the impact of an individual’s sex. These factors may include socioeconomic deprivation, ethnicity, and contact with social services (T. Ford et al., 2007; Madsen et al., 2018; Mennies et al., 2020; Prasad et al., 2019; Wright et al., 2015).

The aim of this study was to use routinely collected, nation-wide primary and secondary healthcare records to examine sex differences in ADHD diagnosis and clinical care of children and young adults diagnosed with ADHD across a 20-year time period. Additional aims were to assess whether females from certain demographic groups experience particular delays in obtaining an ADHD diagnosis and treatment and for changes over time.

## Method

### Data

We used nationwide register data from the Secure Anonymised Information Linkage (SAIL) databank (http://www.saildatabank.com) (Jones et al., 2014; Rodgers et al., 2009, 2012). SAIL is a person-level linkable data repository that covers routinely collected primary and secondary healthcare, education, and social care data for Wales (including approximately 5.45 million individuals). Data were linked using a secure research infrastructure platform, supported by the Adolescent Mental Health Data Platform (ADP: https://adolescentmentalhealth.uk), that pools data from multiple sources to create a research cohort using person-specific anonymous linkage field identity numbers, using all records linked deterministically or probabilistically with a linkage score >0.9 (Lyons et al., 2009). Ethical approval for this project was granted by the Information Governance Review Panel (IGRP #1335); this is an independent body consisting of government, regulatory and professional agencies (D. V. Ford et al., 2009).

The following datasets, covering the whole population of Wales, unless otherwise specified, were linked at individual level. Information on date of birth and sex assigned at birth was obtained from the Welsh Demographic Service Dataset (WDSD), which is a demographics register of people registered with a general practice (GP) in Wales. Data on death was obtained from the Annual District Death Extract (ADDE). The databank includes healthcare records from the Welsh Longitudinal General Practice Database (WLGPD), including attendance and clinical information for primary care data (covering 86% of the population of Wales), the Patient Episode Database for Wales (PEDW), including hospital admissions (inpatients and day cases, including diagnoses), and Emergency Department Dataset (EDDS), including accident and emergency attendance. These datasets include dates of diagnosis/appointments and prescription information.

Information at household level, including the Welsh Index of Multiple Deprivation, was obtained from the WDSD dataset. Information on ethnicity came from the Office for National Statistics (ONS) Census 2011: Welsh Records (CENW), covering 68.9% of the study population. Information on young people who have had contact with social services came from linked databases held by Welsh Government, including the Children Receiving Care and Support (CRCS), Looked After Children Wales (LACW), and Children in Need Wales (CINW) census data (Lee et al., 2022).

### Cohort definition

The study follow-up period used data from 01.01.2000 until 31.12.2019, spanning 20 calendar years. A minimum of 1 year of follow-up time was required and individuals with less than 365 (non-continuous) days of data were excluded. Individuals were included if they were born between 01.01.1989 and 31.12.2013. At the start of the study period, the oldest individuals were 11 years old, allowing a minimum of 1 year of follow-up data before age 12. Each individual was followed up from the maximum date between their 3^rd^ birthday, the start of the study period, start of registration with a GP surgery providing data to SAIL, start of registration at a Welsh address in the WDSD. The end of each individual’s follow-up period was defined as the end of the study period, end of registration with a GP surgery providing data to SAIL, end of registration at a Welsh address, or their date of death (whichever came first). To obtain unbiased estimates of age at first recorded diagnosis and include data from young adults, an individual’s age was not used when defining the end of their follow-up period; by the end of the study period, the oldest individuals were 30 years old.

The sample was defined as those meeting the above inclusion criteria, who received an ADHD diagnosis at any time during each individual’s follow-up period. Individuals who only had a record of ADHD diagnosis prior to age 3 and never again or outside their follow-up period were excluded. ADHD diagnosis was defined using a list of International Classification of Diseases, 10th Revision (ICD-10) codes relevant to ADHD, that are used in secondary care, as well as Read Code lists and algorithms (version 2), which are a coded thesaurus of clinical terms (including diagnoses and administrative codes) used in primary care in the UK National Health Service (NHS). These ADHD codes have previously been validated using an external cohort of young people with ADHD (Langley et al., 2023); see **Table S1**. ADHD medications were not used to define the cohort as they were analysed to examine sex differences.

### Variable definitions

#### Timing of ADHD diagnosis

Age at first recorded ADHD diagnosis was defined using the first date recorded in either primary or secondary care (WLGPD/PEDW) records. Because the precise age of first recorded diagnosis could be influenced by numerous factors, including waiting list variability across geographical location, we also generated binary variables related to diagnosis timing. ‘Earlier’ ADHD diagnoses were defined as those that were first recorded before age 12 years (i.e. prior to or around the time of the key life transition from primary to secondary school and in line with the ADHD symptom age at onset definition used by the DSM-5). ‘Later’ diagnoses were defined as those first recorded on or after an individual’s 12^th^ birthday (i.e. during secondary school or young adulthood). Given the typical early onset of ADHD and its wide-ranging impacts, failure to recognise diagnosis during primary school is likely to indicate a lack of timely educational or other support. We also separately tested diagnosis before and after age 18 (i.e. prior to or around the time of the key life transition to adulthood, including transition from child to adult healthcare services; child: <18, adult >=18).

#### ADHD medication

We defined several variables related to ADHD medication prescriptions, expanding a previous list of Read Codes (Langley et al., 2023) to include additional codes based on consultation with a clinical child psychiatrist specialising in ADHD in Wales (**Table S2**). The following binary variables were defined: prescription of any ADHD medication, prescription of any stimulant medication, prescription of any non-stimulant medication, and prescription of both types of ADHD medication at some time during the study period; this latter variable may represent stimulant treatment failure or intolerance, or alternatively co-occurring tics ((NICE), 2019). We also defined a variable for the time elapsed between the first recorded ADHD diagnosis and ADHD prescription; individuals who were prescribed medication up to 1 month prior to first recorded diagnosis (N=271) were given a score of zero, whereas other negative values (N=2,101) were recoded as missing (these could result from individuals diagnosed before the start of the study).

#### Other neurodevelopmental and mental health conditions

We used lists of Read Codes and ICD-10 codes available in the ADP (see **Table S3** for links to code lists) to derive variables related to recorded instances and ages at first record for a range of conditions. We focused on commonly co-occurring neurodevelopmental conditions: autism spectrum disorder (ASD) and specific learning difficulties (LD) such as dyslexia or motor problems, defined using validated lists of codes (**Table S3**) (John et al., 2022; Langley et al., 2023; Underwood et al., 2022). We defined separate variables for commonly co-occurring mental health conditions: anxiety, depression, and ‘other mental health condition’, including self-harm, eating disorders, alcohol and drug misuse, conduct disorder, bipolar disorder, schizophrenia, and other psychotic disorders (Economou et al., 2012; John et al., 2016, 2018, 2020, 2022; Kennedy et al., 2022; Marchant et al., 2020; Micali et al., 2013; Wood et al., 2019). We also examined the difference between ages at first recorded ADHD and mental health conditions to derive binary variables of whether a mental health condition was recorded prior to ADHD, compared to at the same time or after ADHD.

#### Antidepressant medication

We defined antidepressant medication prescriptions using a previously developed list of Read Codes (**Table S3**) (John et al., 2016). We derived variables for any antidepressant medication prescription, age at first recorded prescription, and a binary variable for whether first recorded antidepressant was prior to or after ADHD diagnosis. To further examine timing of antidepressant use in relation to ADHD diagnosis, we derived a binary variable for whether individuals with an antidepressant prescription recorded prior to first ADHD diagnosis continued to receive this medication after ADHD diagnosis or not.

#### Demographic factors

We examined sex assigned at birth, based on information in the WDSD. Information on ethnicity was available using groupings in line with the major ethnic groups in the UK as defined by the ONS: White, Asian/Asian British, Black/African/Caribbean/Black British, mixed/multiple ethnic groups, and other ethnic minority group; a variable was defined for ethnic majority (White) compared to ethnic minority (all other groups), due to small sample sizes of these groups. Socioeconomic deprivation was defined using the Welsh Index of Multiple Deprivation (WIMD), based on average income, education, employment, and health statistics in a local geographical area, at the start of entry into the study. Quintiles of deprivation were defined using the national WIMD cut-offs (1: least deprived, 5: most deprived). We also defined a subgroup of individuals who have ever had contact with social services, including those who have ever been looked after in state/foster care, based on the LACW dataset, as well as whether they have ever been listed on the child protection register, based on a combination of the CRCS and CINW datasets (Lee et al., 2022). It was not possible to conclusively ascertain from the data which individuals definitely did not have social services contact.

### Data analyses

First, we established the male to female (M:F) sex ratio for the full ADHD cohort. We examined the M:F ratio in individuals stratified by age at first recorded ADHD diagnosis: 1) earlier (<12) vs later (12+) and 2) child (<18) vs adult (18+). We also examined the ratio stratified by birth year (clustering every 5 years to increase cluster sizes), calendar year of first recorded diagnosis (clustering every 5 years), ethnicity, socioeconomic deprivation level, and social services involvement.

Next, we tested for sex differences in variables related to diagnosis and prescriptions for ADHD, other neurodevelopmental conditions (ASD and LD), anxiety, depression, and other mental health conditions. Correction for multiple testing was applied using the false discovery rate (FDR).

Finally, we tested whether similar patterns of associations were seen across different demographic groups. We stratified the analyses by ethnic group, deprivation level, and birth year cluster and used interaction tests to assess whether these variables moderated the observed associations. We also repeated the main analyses in individuals with any social services contact, without doing an interaction test, due to incomplete information for this variable.

All analyses were performed using logistic regression in R, to estimate the odds ratio for each variable, comparing males (coded as 0) and females (coded as 1). Birth year and individual follow-up time were included as covariates, to account for variation in date of birth and length of individual follow-up time.

### Sensitivity analyses

We explored the influence of changing guidelines and services, which could have influenced diagnostic practice in Wales. The ADHD NICE guidelines were published in 2008 and revised in 2018. In April 2016, there was a substantial change in child ADHD diagnostic services in Wales, with the introduction of neurodevelopmental services and a gradual shift over time of diagnosing ADHD in these new services. As such, we checked the M:F ratio stratified by year of first recorded ADHD diagnosis, to explore changes in the sex ratio after 2008 and 2016.

We also tested two other ages to split the sample based on first ADHD diagnosis, corresponding to the transition from UK school key stages 1–2 (age 7) and 3–4 (age 14), to ascertain whether there is a clear point at which the M:F ratio changes and whether this influenced our results.

To consider the influence of diagnostic precision and coverage, we repeated our main analyses in restricted subsets of the cohort. First, we only included data from individuals who were diagnosed with ADHD at least once after age 5, to account for diagnostic uncertainty prior to this age (N=306 excluded) and were also followed up from age 5 onwards, thereby excluding those who entered into the study after their 5^th^ birthday (N=4,157). Finally, we repeated the analyses in individuals with complete or relatively complete data coverage (≥95%) during the time each individual was included in the study (N=9,816).

## Results

### Description of male to female ratios

The cohort consisted of 13,110 (79.7%) males and 3,348 (20.3%) females diagnosed with ADHD, giving a male to female (M:F) ratio of 3.9:1. **Figure 1** displays the M:F ratios (see **Table S4** for details). The M:F ratio was higher in individuals first diagnosed before age 12 compared with after this age, as well as in those diagnosed before age 18 compared to after. The lowest M:F ratio (1.9:1) was seen in those diagnosed at age 18 or above. Correspondingly, the M:F ratio split by birth year clusters showed a lower ratio in those who were oldest by the end of the study period (ages 26–30) and a higher ratio in those who were youngest by the end of the study period (ages 6–10). The M:F ratio also varied by year of diagnosis; those first diagnosed at the beginning of the study period showed the highest M:F ratio (5.1:1) and this ratio decreased over time. **Figure S1** shows the M:F ratio split by individual years, indicating fluctuations but a general downward trend over time, particularly after 2008 when the ADHD NICE guidelines were published. The M:F ratio was similar across ethnicity groups, with a slightly higher ratio in individuals from an ethnic minority compared to ethnic majority background. The M:F ratio was a little lower in individuals who had any contact with social services, compared with the full cohort. The M:F ratio was somewhat higher in those who lived in the most deprived areas.

**Figure 1:**
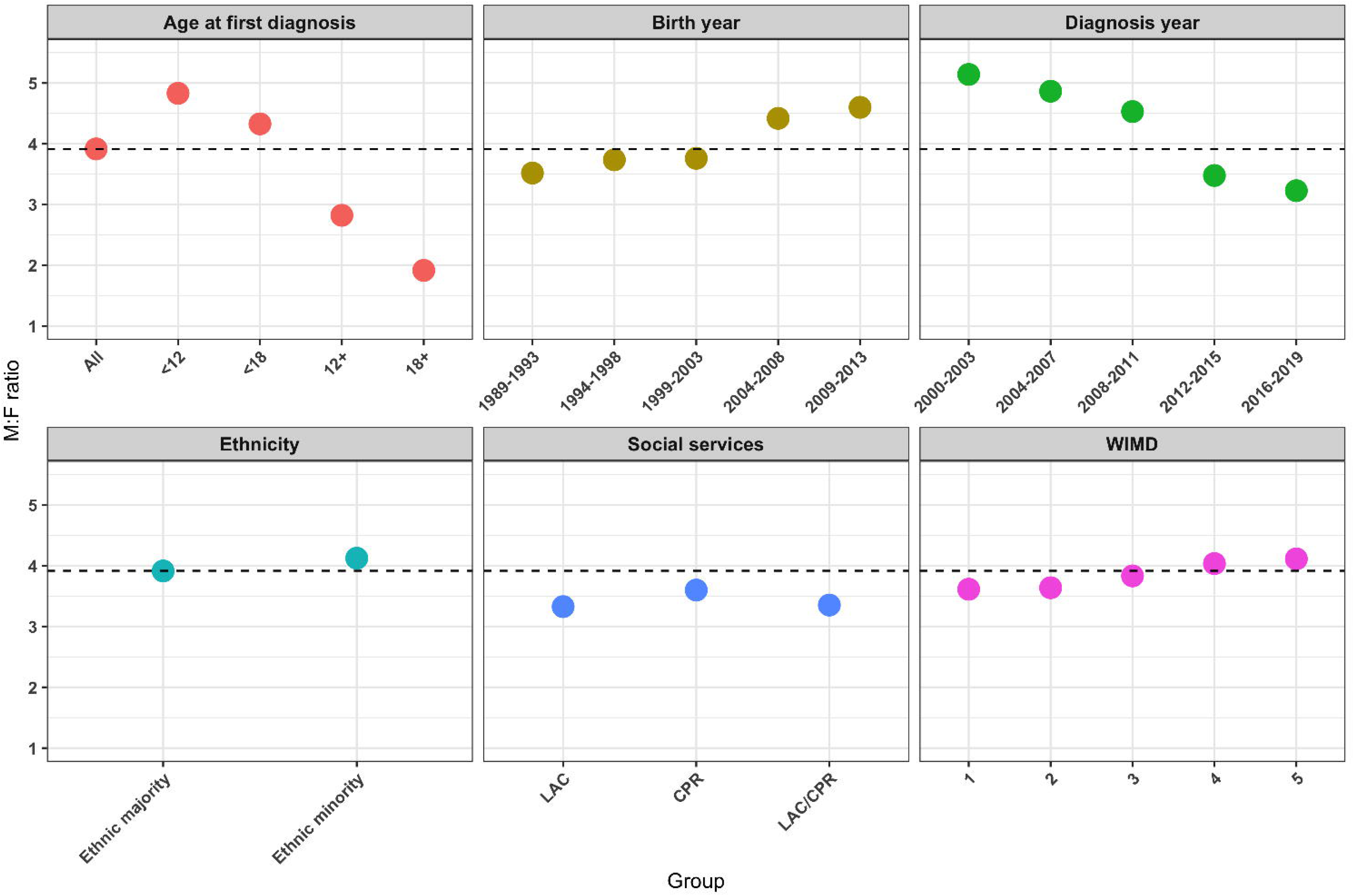
Male to female ratios in the full cohort and stratified by age at first recorded diagnosis, year of birth, year of first recorded diagnosis, ethnicity group, experience of social services involvement, and deprivation level. The dashed line indicates the male to female ratio in the full cohort. LAC: looked after children; CPR: child protection register; WIMD: Welsh Index of Multiple Deprivation (1: least deprived, 5: most deprived).

### Sex differences in ADHD diagnosis and clinical care

The results of analyses examining sex differences in ADHD diagnosis and clinical care can be found in **Figure 2** and **Table S5** (detailed results).

**Figure 2:**
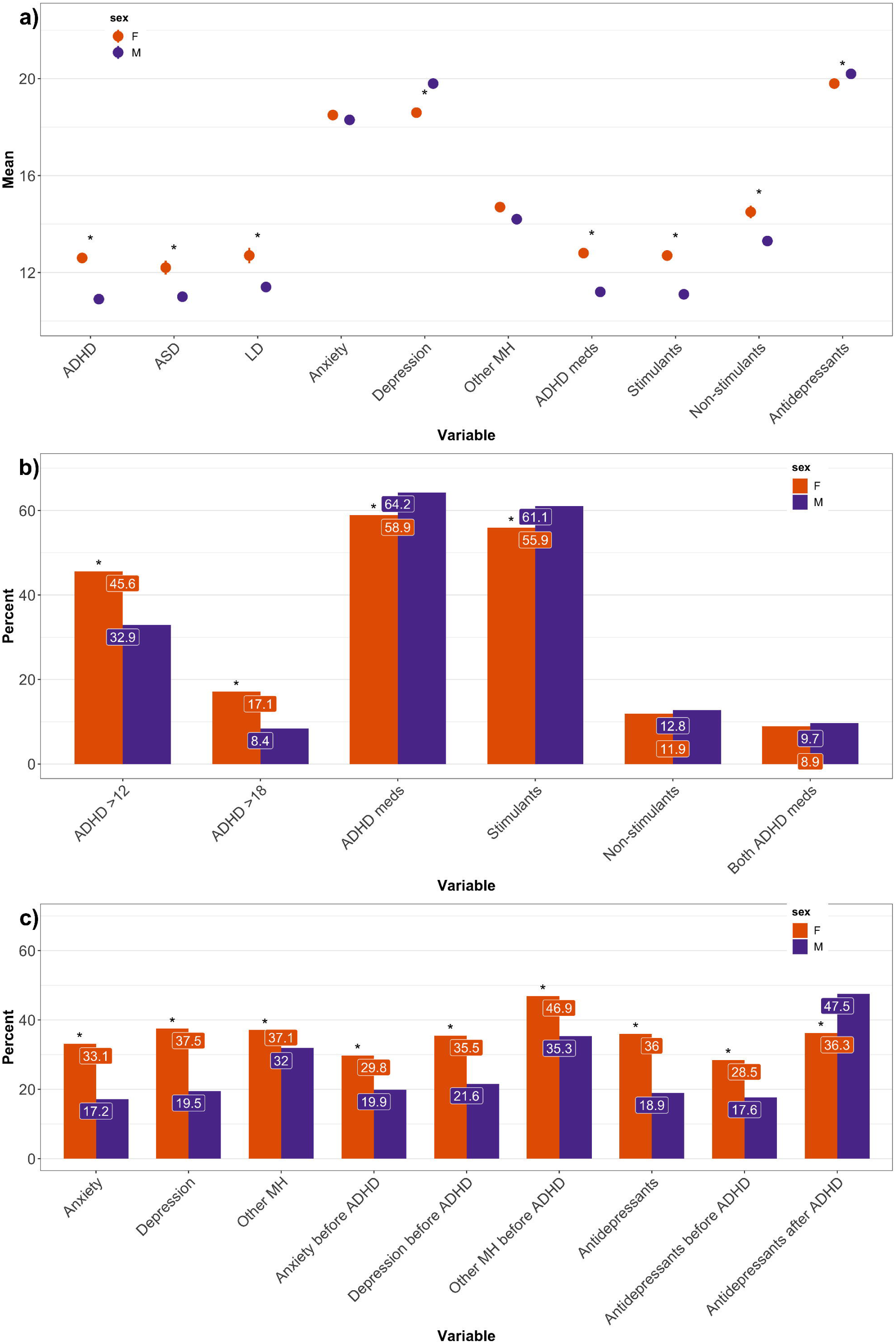
Sex-stratified results displaying: a) ages at first recorded instances of diagnoses and prescriptions (with standard errors of the mean shown as error bars), b) percentages related to recorded ADHD diagnosis timing and ADHD prescriptions, and c) percentages related to recorded mental health diagnoses and antidepressant prescriptions. Where stated “before ADHD” in Figure 2c, this variable is measured only within those who have a record of the specified diagnosis/prescription. Where stated “after ADHD” in Figure 2c, this variable is measured only within those who have a recorded antidepressant prescription prior to ADHD diagnosis. ASD: autism spectrum disorder; LD: learning difficulties; MH: mental health. * indicates p<0.05 after false discovery rate correction.

#### Timing of ADHD diagnosis

On average, males were first diagnosed with ADHD at a younger age than females (M: 10.9, F: 12.6 years; **Figure 2a**). Females were more likely than males to be diagnosed after age 12 and also after age 18 (**Figure 2b**).

#### ADHD medication

Males were more likely to be prescribed any ADHD medication and specifically stimulant medication, with no sex difference in non-stimulant prescriptions or being prescribed both types of medications (**Figure 2b**). Additionally, males were younger at first prescription of ADHD medications (M: 11.2, F: 12.8 years), including for both stimulants (M: 11.1, F: 12.7 years) and non-stimulants separately (M: 13.3, F: 14.5 years; **Figure 2a**). There was no sex difference in the time elapsed between first recorded ADHD diagnosis and first prescribed medication for ADHD (M: 1.1, F: 1.1 years; **Table S5**).

#### Other neurodevelopmental and mental health conditions

Males were younger at first recorded diagnoses of co-occurring ASD (M: 11.0, F: 12.2 years) and LD (M: 11.4, F: 12.7 years; **Figure 2a**). Females were more likely to have a recorded diagnosis of anxiety, depression, or any other mental health condition, in addition to ADHD (**Figure 2c**). Females were younger at first recorded depression (F: 18.6, M: 19.8 years), with no sex difference at age of first anxiety (F: 18.5, M: 18.3 years) or other mental health conditions (F: 14.7, M: 14.2 years; **Figure 2a**). In those with the co-occurring condition, females were more likely than males to have a record of each of the co-occurring diagnoses prior to first recorded ADHD diagnosis (**Figure 2c**).

#### Antidepressant medication

Females were more likely to be prescribed antidepressant medication (**Figure 2c**), were younger at first antidepressant prescription (F: 19.8, M: 20.2 years; **Figure 2a**), and were more likely to be prescribed antidepressant medication prior to their ADHD diagnosis (**Figure 2c**). In the subgroup of individuals who received antidepressant medication prior to ADHD diagnosis, females were less likely than males to continue to receive antidepressants after their recorded ADHD diagnosis (**Figure 2c**).

### Impact of demographic factors

We repeated the analyses in the subset of individuals who had ever had social services involvement (N=1,332). Males were on average diagnosed at a younger age than females (M: 9.6, F: 10.7 years), with mean ages somewhat lower than seen in the full cohort. Overall, the pattern of sex differences was similar to the primary analyses (**Table S6**), except the association was no longer robust for some variables (ages at non-stimulant and antidepressant medications and LD diagnosis; anxiety and antidepressant records before ADHD, and antidepressant continuation), likely due to the reduced sample size as the point estimates were mostly similar (except for anxiety before ADHD).

Next, we stratified the analyses by ethnicity, WIMD, and birth year, and tested for moderation of sex differences by these variables. **Table 1** summarises the results of the interaction analyses. There was no evidence of moderation from ethnicity or WIMD on any of the variables.

**Table 1:**
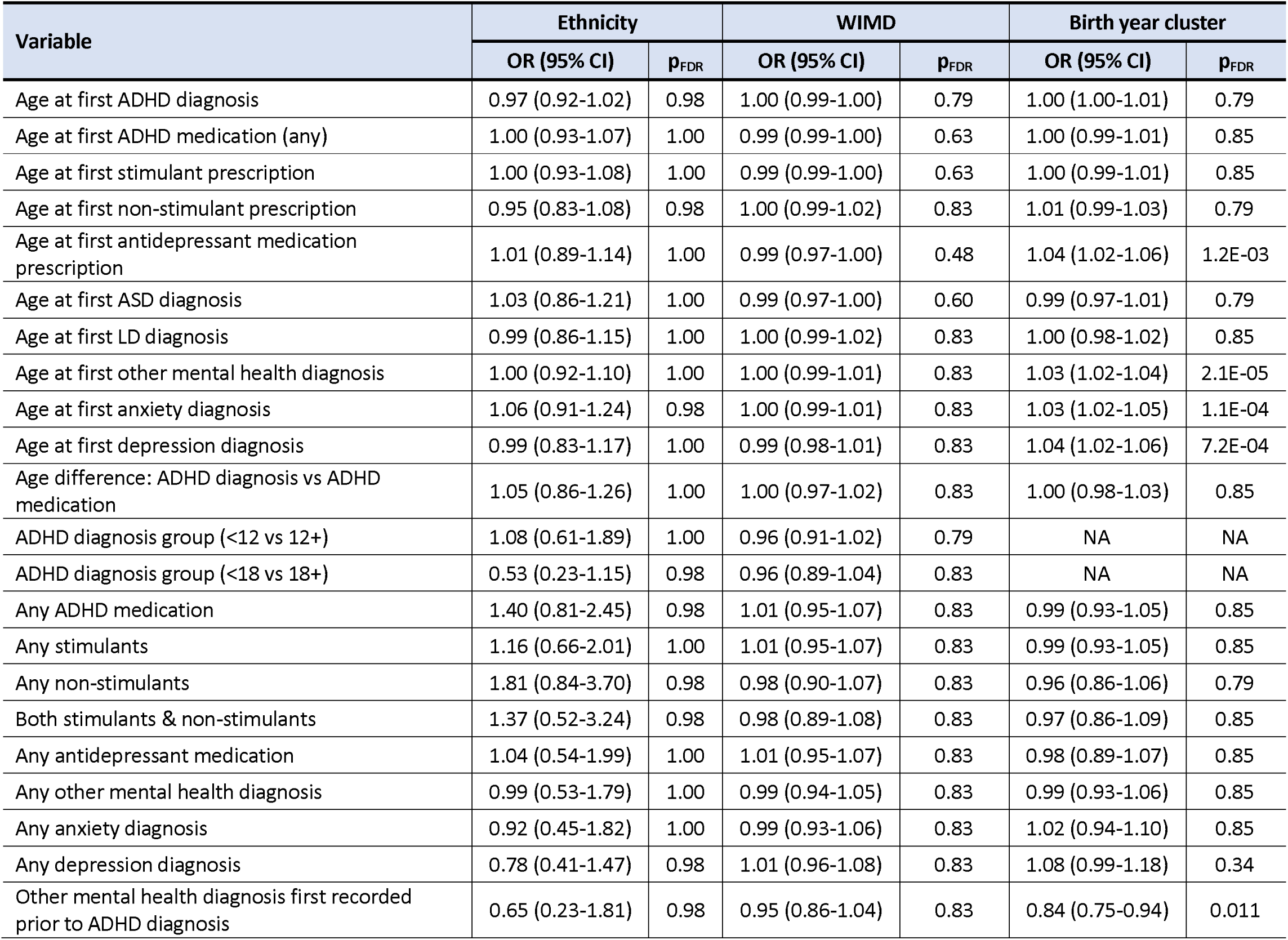
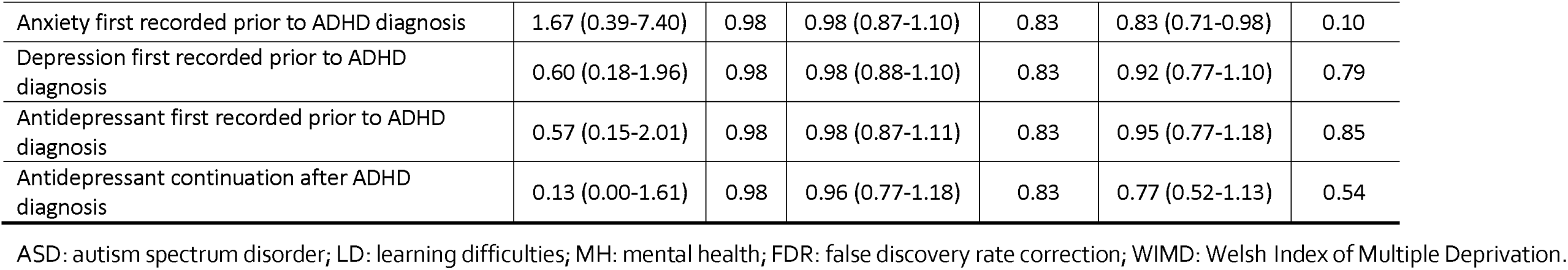
Influence of demographic variables on sex differences in ADHD diagnosis and clinical care.

| Variable | Ethnicity |  | WIMD |  | Birth year cluster |  |
| --- | --- | --- | --- | --- | --- | --- |
|  | OR (95% CI) | p<sub>FDR</sub> | OR (95% CI) | p<sub>FDR</sub> | OR (95% CI) | p<sub>FDR</sub> |
| Age at first ADHD diagnosis | 0.97 (0.92-1.02) | 0.98 | 1.00 (0.99-1.00) | 0.79 | 1.00 (1.00-1.01) | 0.79 |
| Age at first ADHD medication (any) | 1.00 (0.93-1.07) | 1.00 | 0.99 (0.99-1.00) | 0.63 | 1.00 (0.99-1.01) | 0.85 |
| Age at first stimulant prescription | 1.00 (0.93-1.08) | 1.00 | 0.99 (0.99-1.00) | 0.63 | 1.00 (0.99-1.01) | 0.85 |
| Age at first non-stimulant prescription | 0.95 (0.83-1.08) | 0.98 | 1.00 (0.99-1.02) | 0.83 | 1.01 (0.99-1.03) | 0.79 |
| Age at first antidepressant medication prescription | 1.01 (0.89-1.14) | 1.00 | 0.99 (0.97-1.00) | 0.48 | 1.04 (1.02-1.06) | 1.2E-03 |
| Age at first ASD diagnosis | 1.03 (0.86-1.21) | 1.00 | 0.99 (0.97-1.00) | 0.60 | 0.99 (0.97-1.01) | 0.79 |
| Age at first LD diagnosis | 0.99 (0.86-1.15) | 1.00 | 1.00 (0.99-1.02) | 0.83 | 1.00 (0.98-1.02) | 0.85 |
| Age at first other mental health diagnosis | 1.00 (0.92-1.10) | 1.00 | 1.00 (0.99-1.01) | 0.83 | 1.03 (1.02-1.04) | 2.1E-05 |
| Age at first anxiety diagnosis | 1.06 (0.91-1.24) | 0.98 | 1.00 (0.99-1.01) | 0.83 | 1.03 (1.02-1.05) | 1.1E-04 |
| Age at first depression diagnosis | 0.99 (0.83-1.17) | 1.00 | 0.99 (0.98-1.01) | 0.83 | 1.04 (1.02-1.06) | 7.2E-04 |
| Age difference: ADHD diagnosis vs ADHD medication | 1.05 (0.86-1.26) | 1.00 | 1.00 (0.97-1.02) | 0.83 | 1.00 (0.98-1.03) | 0.85 |
| ADHD diagnosis group (<12 vs 12+) | 1.08 (0.61-1.89) | 1.00 | 0.96 (0.91-1.02) | 0.79 | NA | NA |
| ADHD diagnosis group (<18 vs 18+) | 0.53 (0.23-1.15) | 0.98 | 0.96 (0.89-1.04) | 0.83 | NA | NA |
| Any ADHD medication | 1.40 (0.81-2.45) | 0.98 | 1.01 (0.95-1.07) | 0.83 | 0.99 (0.93-1.05) | 0.85 |
| Any stimulants | 1.16 (0.66-2.01) | 1.00 | 1.01 (0.95-1.07) | 0.83 | 0.99 (0.93-1.05) | 0.85 |
| Any non-stimulants | 1.81 (0.84-3.70) | 0.98 | 0.98 (0.90-1.07) | 0.83 | 0.96 (0.86-1.06) | 0.79 |
| Both stimulants & non-stimulants | 1.37 (0.52-3.24) | 0.98 | 0.98 (0.89-1.08) | 0.83 | 0.97 (0.86-1.09) | 0.85 |
| Any antidepressant medication | 1.04 (0.54-1.99) | 1.00 | 1.01 (0.95-1.07) | 0.83 | 0.98 (0.89-1.07) | 0.85 |
| Any other mental health diagnosis | 0.99 (0.53-1.79) | 1.00 | 0.99 (0.94-1.05) | 0.83 | 0.99 (0.93-1.06) | 0.85 |
| Any anxiety diagnosis | 0.92 (0.45-1.82) | 1.00 | 0.99 (0.93-1.06) | 0.83 | 1.02 (0.94-1.10) | 0.85 |
| Any depression diagnosis | 0.78 (0.41-1.47) | 0.98 | 1.01 (0.96-1.08) | 0.83 | 1.08 (0.99-1.18) | 0.34 |
| Other mental health diagnosis first recorded prior to ADHD diagnosis | 0.65 (0.23-1.81) | 0.98 | 0.95 (0.86-1.04) | 0.83 | 0.84 (0.75-0.94) | 0.011 |
| Anxiety first recorded prior to ADHD diagnosis | 1.67 (0.39-7.40) | 0.98 | 0.98 (0.87-1.10) | 0.83 | 0.83 (0.71-0.98) | 0.10 |
| Depression first recorded prior to ADHD diagnosis | 0.60 (0.18-1.96) | 0.98 | 0.98 (0.88-1.10) | 0.83 | 0.92 (0.77-1.10) | 0.79 |
| Antidepressant first recorded prior to ADHD diagnosis | 0.57 (0.15-2.01) | 0.98 | 0.98 (0.87-1.11) | 0.83 | 0.95 (0.77-1.18) | 0.85 |
| Antidepressant continuation after ADHD diagnosis | 0.13 (0.00-1.61) | 0.98 | 0.96 (0.77-1.18) | 0.83 | 0.77 (0.52-1.13) | 0.54 |

ASD: autism spectrum disorder; LD: learning difficulties; MH: mental health; FDR: false discovery rate correction; WIMD: Welsh Index of Multiple Deprivation.

Birth year had an influence on several variables related to mental health diagnoses and antidepressant medication. Overall, females were more likely to obtain an ‘other’ mental health diagnosis prior to ADHD, but only in the individuals in the three earlier birth year clusters. Females were younger than males at first recorded antidepressant medication, but this was only observed in the two earlier birth year clusters. Females were younger at first depression diagnosis compared to males, but only in those born in the three earlier birth year clusters. Finally, females were older at first anxiety and other mental health diagnoses compared to males, but only in those born between 1999-2008 and not in the other birth year clusters. Birth year was incorporated into all other analyses as a covariate, adjusting for these effects.

### Sensitivity analyses

When using ages 7 and 14 to split the sample into age at ADHD diagnosis groups, there was a similar pattern of results. The M:F ratio was higher in those diagnosed before age 7 (5.1:1) or age 14 (4.7:1) compared with those diagnosed at age 7 onwards (3.7:1) or age 14 onwards (2.3:1), respectively. Females were more likely than males to be in the later diagnosed group (age 7: 85.9% vs 81.6%, OR(CIs)=1.30(1.16-1.45); age 14: 34.2% vs 20.2%, OR(CIs)=2.18(1.98-2.39)). Overall, the results showed gradually increasing strengths of association for this analysis across ages 7, 12, 14 and 18.

Restricting the sample to those with ADHD diagnoses after age 5 and those with less missing information (N=12,301; 74.7%) showed a largely similar pattern of results, except that in this subgroup, females were older at first recorded anxiety or other mental health diagnosis and were less likely to have been prescribed non-stimulants as well as both types of ADHD medications; on the other hand the sex difference for age at first antidepressant medication no longer held (**Table S7**). Restricting the sample to those with good coverage across the study period (N=9,816; 59.6%) no longer showed a sex difference for age at first antidepressant medication or first recorded LD, but did show the sex difference for ages at first recorded anxiety or other mental health diagnosis (**Table S8**).

## Discussion

The aim of this study was to test for sex differences in diagnosis and clinical care of ADHD in children and young adults (aged 3–30 years), particularly regarding recognition and treatment of other mental health conditions, using national healthcare records. Females were older at age of first recorded diagnoses of ADHD and co-occurring ASD/LD and less likely to be prescribed stimulant medication. They were more likely to be diagnosed with anxiety, depression, or another mental health condition and to be prescribed antidepressant medication, both in general and prior to their ADHD diagnosis. They were younger at first antidepressant prescription and less likely to continue to receive antidepressants after ADHD diagnosis. These sex differences in diagnosis and treatment of young people with ADHD have important implications for improvements to clinical practice.

Overall, there were almost 4 males for every female diagnosed with ADHD, similar to other studies (Sayal et al., 2018; Willcutt, 2012). Regardless of which sub-strata of the cohort was examined, the M:F ratio was always above 1. The lowest ratio was in individuals diagnosed in adulthood (1.9:1). The other factor influencing the ratio was year of diagnosis, with a decrease over time (from 5.1:1 to 3.2:1). While the absolute numbers of both sexes being diagnosed increased over time (**Table S4**), this increase appears more rapid in females; however in later years, individuals in the cohort are older, which may explain this. Ethnicity and social services involvement showed only a small influence, indicating that these factors do not exacerbate under-recognition of female ADHD. However, females from lower socioeconomic backgrounds may be less likely to be recognised, referred, and diagnosed than those from higher socioeconomic status, although the difference was small (4.1:1 vs 3.6:1).

Consistent with Scandinavian research, females were older at first ADHD diagnosis (Dalsgaard et al., 2019; Martin et al., 2018; Wimberley et al., 2020). ADHD diagnosis requires symptom onset before age 12 (DSM-5 and ICD-11). We observed that approximately a third of males (32.9%) and nearly half of females (45.6%) were diagnosed after age 12 and a substantial subgroup (8.4% males, 17.1% females) were diagnosed in adulthood. Later age at diagnosis in females was also seen for co-occurring neurodevelopmental conditions, indicating that a sex bias is even seen in individuals with likely more complex needs and difficulties.

Our study extends previous findings that females are less likely to be prescribed ADHD medication (Dalsgaard et al., 2014; Kok et al., 2020; Russell et al., 2019), by considering stimulants and non-stimulants separately. We found that females were less likely to be prescribed stimulants but with no sex difference for non-stimulants or being prescribed both medication types, though the sensitivity analyses did suggest lower rates of non-stimulant prescription and being prescribed both medication types in females as well. Although females were older at first ADHD medication prescription (regardless of type), later treatment could be explained by later diagnosis. It is unclear why females are less likely to be prescribed medication. Although they may have lower symptom severity (Loyer Carbonneau et al., 2021), sex differences in prescriptions exist even when accounting for severity (Russell et al., 2019).

It has been proposed that ADHD in females may be missed due to diagnostic overshadowing from mental health conditions (Quinn & Madhoo, 2014; Young et al., 2020). Our study provides some of the first robust empirical support for this, by finding that females are not only more likely to be recognised as having a range of other mental health difficulties, but also that these conditions were more likely to have been diagnosed prior to the recognition of ADHD in females. Depression was diagnosed earlier in females. There was no sex difference in age at diagnosis of other mental health conditions, except in sensitivity analyses, which showed the opposite effect. Females were also more likely to be prescribed antidepressant medication and at a younger age than males. This was more likely to be prior to ADHD diagnosis and in those prescribed antidepressants prior to ADHD diagnosis, females were less likely to continue to receive antidepressant prescriptions after ADHD diagnosis. Consistent with these findings, previous research indicated that anxiety diagnosis may precede ADHD diagnosis in females, clinical referral was more likely to be for emotional problems in females, and females were more likely to receive non-ADHD medication and have been admitted to psychiatric inpatient care (Klefsjö et al., 2021; Martin et al., 2020).

The results of our study indicate that anxiety, depression, and other mental health difficulties are more likely to be recognised prior to ADHD in females, leading to referral to specialist services. These conditions may contribute to diagnostic overshadowing, with ADHD only recognised and diagnosed at a later age. The fact that antidepressant treatment is more likely to be discontinued in females after diagnosis suggests that this treatment may be no longer considered necessary once underlying ADHD is recognised. This further suggests that for some females, there was initial misdiagnosis with another condition prior to ADHD diagnosis.

We found limited evidence that social services involvement, ethnic minority status, and socioeconomic deprivation intersect with sex to influence ADHD clinical care, although these factors play a primary role in referral and recognition of ADHD (T. Ford et al., 2007; Madsen et al., 2018; Mennies et al., 2020; Prasad et al., 2019; Wright et al., 2015). One limitation is that the sample sizes for some subgroups (e.g. ethnic minority) were relatively small, which reduced our power to detect differences. We were also unable to examine more specific ethnic groups due to small sample sizes and incomplete data. Our study focused on young people diagnosed with ADHD and did not examine differences relative to the general population. We were also not able to examine non-pharmacological interventions.

Despite these limitations, our study had several strengths, including a large nation-wide ADHD cohort and use of real-world primary and secondary healthcare records, thereby providing a representative picture of clinical practice in Wales during the study period. We examined M:F ratios over time and found a generally decreasing trend. Although clinical services have changed over time (e.g. new NICE guidelines and neurodevelopmental services), there have also been influences from mainstream and social media and other factors (e.g. changes in professional training), which have increased awareness and likely improved recognition of female ADHD.

## Conclusion

Overall, we observed that while relatively more females are being diagnosed with ADHD over time, there is still a sex bias and there are prominent sex differences in provision and timing of diagnosis and treatment for ADHD and co-occurring conditions. Our study adds to the growing evidence base that females with ADHD are experiencing barriers in timely access to clinical care and treatment, which may be partly explained by diagnostic overshadowing from anxiety, depression, and other mental health difficulties or initial misdiagnosis. Further efforts to improve timely recognition of female ADHD by clinicians and referrers (e.g. teachers and parents) are required.

### Key points and relevance

- Previous research suggests that females are less likely to be diagnosed with ADHD, are diagnosed later, and are less likely to be prescribed stimulant ADHD medication.
- This study observed that females with ADHD are diagnosed later even in the presence of cooccurring neurodevelopmental conditions and are more likely to be diagnosed with anxiety, depression or other mental health conditions and be prescribed antidepressants, particularly prior to their ADHD diagnosis.
- Females are also less likely to continue to receive antidepressant medication after ADHD diagnosis.
- Clinical services need to improve timely access to clinical care, recognition, and treatment for females with ADHD, with clinicians being vigilant about possible diagnostic overshadowing from other mental health difficulties and initial misdiagnosis.

## Data Availability

Data are available through application to the SAIL databank.

http://www.saildatabank.com

## Acknowledgements

This study was funded by the Welsh Government through Health and Care Research Wales via a National Institute for Health and Care Research (NIHR) Advanced Fellowship (Ref: NIHR-FS(A)-2022) and was also supported by a NARSAD Young Investigator Grant from the Brain & Behavior Research Foundation (grant no. 27879).

KS and TF are NIHR Senior Investigators. The views expressed are those of the authors and not necessarily those of the NIHR or the Department of Health and Social Care.

This work was supported by the Adolescent Mental Health Data Platform (ADP). The ADP is funded by MQ Mental Health Research Charity (Grant Reference MQBF/3 ADP). The views expressed are entirely those of the authors and should not be assumed to be the same as those of ADP or MQ Mental Health Research Charity.

This study makes use of anonymised data held in the Secure Anonymised Information Linkage (SAIL) Databank. We would like to acknowledge all the data providers who make anonymised data available for research.

The study also makes use of data provided by the Office of National Statistics (ONS) and we acknowledge that those who carried out the original collection and analysis of the ONS data bear no responsibility for the further analysis or interpretation.

This work was supported by the Wolfson Centre for Young People’s Mental Health, established with support from the Wolfson Foundation.

## Correspondence

Joanna Martin (Mailing address: Centre for Neuropsychiatric Genetics and Genomics, Cardiff University, Hadyn Ellis Building, Maindy Road, Cardiff CF24 4HQ, UK).

## Access to data and data sharing

JM and OR confirm that they had full access to all the data in the study and take responsibility for the integrity of the data and the accuracy of the data analysis. Data are available through application to the SAIL databank (http://www.saildatabank.com).

