## Supplement for "Sex differences in ADHD diagnosis and clinical care: A national study of population healthcare records in Wales"

### Table S1: Attention deficit hyperactivity disorder (ADHD) diagnosis definition: ICD-10 codes used in secondary care (hospital and emergency services) & Read Codes used in primary care

| **Code** | **Description** |
| --- | --- |
| F90 | ICD-10 hyperkinetic disorders |
| F90.0 | ICD-10 disturbance of activity and attention |
| F90.1 | ICD-10 hyperkinetic conduct disorder |
| F90.2 | ICD-10 attention-deficit hyperactivity disorder, combined type |
| F90.8 | ICD-10 other hyperkinetic disorders |
| F90.9 | ICD-10 hyperkinetic disorder, unspecified |
| 6A61. | adhd annual review \| attention deficit hyperactivity disorder annual review |
| 8BPT. | drug therapy for adhd \| drug therapy adhd (attention deficit hyperactivity disorder) \| drug therapy for adhd (attention deficit hyperactivity disorder) |
| 8BPT0 | stimulant drug therapy adhd \| stimulant drug therapy for adhd \| stimulant drug therapy for adhd (attention deficit hyperactivity disorder) |
| 8BPT1 | non-stimulnt drug therapy adhd \| non-stimulant drug therapy for adhd \| non-stimulant drug therapy for adhd (attention deficit hyperactivity disorder) |
| 9Ngp. | on drug therapy adhd \| on drug therapy adhd (attention deficit hyperactivity disorder) \| on drug therapy for adhd (attention deficit hyperactivity disorder) |
| 9Ngp0 | on stimulant drug therapy adhd \| on stim drug therapy adhd (attention def hyperactivity disordr) \| on stimulant drug therapy for adhd (attention deficit hyperactivity disorder) |
| 9Ngp1 | on non-stimulnt drug therapy adhd \| on non-stimulant drug therapy for adhd \| on non-stimulant drug therapy for adhd (attention deficit hyperactivity disorder) |
| E2E.. | childhood hyperkinetic syndr. \| childhood hyperkinetic syndrome |
| E2E0. | child attention deficit disord \| child attention deficit disorder |
| E2E00 | attention deficit-not hyperact \| attention deficit without hyperactivity |
| E2E01 | attention deficit +hyperactive \| attention deficit with hyperactivity |
| E2E0z | child attent.deficit dis.nos \| child attention deficit disorder nos |
| E2E1. | hyperkinesis+development delay \| hyperkinesis with developmental delay |
| E2E2. | hyperkinetic conduct disorder |
| E2Ey. | other hyperkinetic manifestat. \| other hyperkinetic manifestation |
| E2Ez. | hyperkinetic syndrome nos |
| Eu90. | [x]hyperkinetic disorders |
| Eu900 | [x]disturbance activity/attntn \| [x]disturbance of activity and attention |
| Eu901 | [x]hyperkinetic conduct disord \| [x]hyperkinetic conduct disorder |
| Eu9y7 | [x]attention deficit disorder |
| ZS91. | attention deficil disorder |

### Table S2: ADHD prescriptions used in primary care

| **Code** | **Description** | **Stimulant / non-stimulant?** |
| --- | --- | --- |
| dc1.. | DEXAMFETAMINE SULFATE | stimulant |
| dc11. | *DEXEDRINE 5mg tablets | stimulant |
| dc1v. | DEXAMFET SULF 1mg/mL oral soln | stimulant |
| dc1w. | DEXAMFETAMINE SULPH 5mg tabs | stimulant |
| dc1x. | *DEXAMPHETAMINE 7.5mg m/r caps | stimulant |
| dc1y. | *DEXAMPHETAMINE 12.5mg caps | stimulant |
| dc1z. | *DEXAMPHETAMINE 20mg m/r caps | stimulant |
| dw1.. | METHYLPHENIDATE | stimulant |
| dw11. | METHYLPHENIDATE HCL 10mg tabs | stimulant |
| dw12. | RITALIN 10mg tablets | stimulant |
| dw13. | *EQUASYM 5mg tablets | stimulant |
| dw14. | *EQUASYM 20mg tablets | stimulant |
| dw15. | *EQUASYM 10mg tablets | stimulant |
| dw16. | EQUASYM XL 20mg m/r capsules | stimulant |
| dw17. | CONCERTA XL 18mg m/r tablets | stimulant |
| dw18. | CONCERTA XL 36mg m/r tablets | stimulant |
| dw19. | *TRANQUILYN 5mg tablets | stimulant |
| dw1A. | *TRANQUILYN 10mg tablets | stimulant |
| dw1B. | *TRANQUILYN 20mg tablets | stimulant |
| dw1C. | EQUASYM XL 10mg m/r capsules | stimulant |
| dw1D. | EQUASYM XL 30mg m/r capsules | stimulant |
| dw1E. | MEDIKINET XL 10mg m/r capsules | stimulant |
| dw1F. | MEDIKINET XL 20mg m/r capsules | stimulant |
| dw1G. | MEDIKINET XL 30mg m/r capsules | stimulant |
| dw1H. | MEDIKINET XL 40mg m/r capsules | stimulant |
| dw1I. | CONCERTA XL 27mg m/r tablets | stimulant |
| dw1J. | MEDIKINET 5mg tablets | stimulant |
| dw1K. | MEDIKINET 10mg tablets | stimulant |
| dw1L. | MEDIKINET 20mg tablets | stimulant |
| dw1M. | MEDIKINET XL 5mg m/r capsules | stimulant |
| dw1N. | MEDIKINET XL 50mg m/r capsules | stimulant |
| dw1O. | MEDIKINET XL 60mg m/r capsules | stimulant |
| dw1P. | MATORIDE XL 18mg m/r tablets | stimulant |
| dw1Q. | MATORIDE XL 36mg m/r tablets | stimulant |
| dw1R. | MATORIDE XL 54mg m/r tablets | stimulant |
| dw1S. | XENIDATE XL 18mg m/r tablets | stimulant |
| dw1T. | XENIDATE XL 36mg m/r tablets | stimulant |
| dw1U. | CONCERTA XL 54mg m/r tablets | stimulant |
| dw1n. | METHYLPHENIDATE 54mg m/r tabs | stimulant |
| dw1o. | METHYLPHENIDATE 50mg m/r caps | stimulant |
| dw1p. | METHYLPHENIDATE 60mg m/r caps | stimulant |
| dw1q. | METHYLPHENIDATE 5mg m/r caps | stimulant |
| dw1r. | METHYLPHENIDATE 27mg m/r tabs | stimulant |
| dw1s. | METHYLPHENIDATE 40mg m/r caps | stimulant |
| dw1t. | METHYLPHENIDATE 10mg m/r caps | stimulant |
| dw1u. | METHYLPHENIDATE 30mg m/r caps | stimulant |
| dw1v. | METHYLPHENIDATE 36mg m/r tabs | stimulant |
| dw1w. | METHYLPHENIDATE 18mg m/r tabs | stimulant |
| dw1x. | METHYLPHENIDATE 20mg m/r caps | stimulant |
| dw1y. | METHYLPHENIDATE HCL 5mg tabs | stimulant |
| dw1z. | METHYLPHENIDATE HCL 20mg tabs | stimulant |
| dw2.. | ATOMOXETINE | non-stimulant |
| dw21. | STRATTERA 10mg capsules | non-stimulant |
| dw22. | STRATTERA 18mg capsules | non-stimulant |
| dw23. | STRATTERA 25mg capsules | non-stimulant |
| dw24. | STRATTERA 40mg capsules | non-stimulant |
| dw25. | STRATTERA 60mg capsules | non-stimulant |
| dw26. | STRATTERA 80mg capsules | non-stimulant |
| dw27. | STRATTERA 100mg capsules | non-stimulant |
| dw28. | STRATTERA 4mg/mL oral solution | non-stimulant |
| dw2s. | ATOMOXETINE 4mg/mL oral soln | non-stimulant |
| dw2t. | ATOMOXETINE 100mg capsules | non-stimulant |
| dw2u. | ATOMOXETINE 80mg capsules | non-stimulant |
| dw2v. | ATOMOXETINE 60mg capsules | non-stimulant |
| dw2w. | ATOMOXETINE 40mg capsules | non-stimulant |
| dw2x. | ATOMOXETINE 25mg capsules | non-stimulant |
| dw2y. | ATOMOXETINE 18mg capsules | non-stimulant |
| dw2z. | ATOMOXETINE 10mg capsules | non-stimulant |
| dw3.. | LISDEXAMFETAMINE | stimulant |
| dw31. | ELVANSE 30mg capsules | stimulant |
| dw32. | ELVANSE 50mg capsules | stimulant |
| dw33. | ELVANSE 70mg capsules | stimulant |
| dw34. | ELVANSE ADULT 30mg capsules | stimulant |
| dw35. | ELVANSE ADULT 50mg capsules | stimulant |
| dw36. | ELVANSE ADULT 70mg capsules | stimulant |
| dw37. | ELVANSE 20mg capsules | stimulant |
| dw38. | ELVANSE 40mg capsules | stimulant |
| dw39. | ELVANSE 60mg capsules | stimulant |
| dw3u. | LISDEXAMFETAMINE DIM 60mg caps | stimulant |
| dw3v. | LISDEXAMFETAMINE DIM 40mg caps | stimulant |
| dw3w. | LISDEXAMFETAMINE DIM 20mg caps | stimulant |
| dw3x. | LISDEXAMFETAMINE DIM 70mg caps | stimulant |
| dw3y. | LISDEXAMFETAMINE DIM 50mg caps | stimulant |
| dw3z. | LISDEXAMFETAMINE DIM 30mg caps | stimulant |
| dw4.. | GUANFACINE | non-stimulant |
| dw41. | INTUNIV 1mg m/r tablets | non-stimulant |
| dw42. | GUANFACINE 1mg m/r tablets | non-stimulant |
| dw43. | INTUNIV 2mg m/r tablets | non-stimulant |
| dw44. | GUANFACINE 2mg m/r tablets | non-stimulant |
| dw45. | INTUNIV 3mg m/r tablets | non-stimulant |
| dw46. | GUANFACINE 3mg m/r tablets | non-stimulant |
| dw47. | INTUNIV 4mg m/r tablets | non-stimulant |
| dw48. | GUANFACINE 4mg m/r tablets | non-stimulant |

### Table S3: Validated code lists for other neurodevelopmental and mental health conditions and prescriptions

| **Name** | **Source** | **Link to concept library code list(s)** |
| --- | --- | --- |
| Anxiety | WLGP, EDDS & PEDW | <https://conceptlibrary.saildatabank.com/phenotypes/PH1113/detail/> |
| Depression | WLGP, EDDS & PEDW | <https://conceptlibrary.saildatabank.com/phenotypes/PH1114/detail/> |
| Self-harm | WLGP, EDDS & PEDW | <https://conceptlibrary.saildatabank.com/phenotypes/PH936/detail/> |
| Autism spectrum disorder | WLGP, EDDS & PEDW | <https://conceptlibrary.saildatabank.com/phenotypes/PH933/detail/> |
| Alcohol misuse | WLGP, EDDS & PEDW | <https://conceptlibrary.saildatabank.com/phenotypes/PH1107/detail/> |
| Drugs misuse | WLGP, EDDS & PEDW | <https://conceptlibrary.saildatabank.com/phenotypes/PH1108/detail/> |
| Schizophrenia | WLGP, EDDS & PEDW | <https://conceptlibrary.saildatabank.com/concepts/C2716/detail/>; <https://conceptlibrary.saildatabank.com/concepts/C2939/detail/> |
| Bipolar disorder | WLGP, EDDS & PEDW | <https://conceptlibrary.saildatabank.com/concepts/C2714/detail/>; <https://conceptlibrary.saildatabank.com/concepts/C2932/detail/> |
| Eating disorder | WLGP, EDDS & PEDW | <https://conceptlibrary.saildatabank.com/phenotypes/PH1116/detail/> |
| Learning difficulties | WLGP, EDDS & PEDW | <https://conceptlibrary.saildatabank.com/phenotypes/PH935/detail/> |
| Conduct disorder | WLGP, EDDS & PEDW | <https://conceptlibrary.saildatabank.com/phenotypes/PH934/detail/> |
| Other psychotic disorder | WLGP, EDDS & PEDW | <https://conceptlibrary.saildatabank.com/concepts/C3160/detail/>; <https://conceptlibrary.saildatabank.com/concepts/C3159/detail/> |
| Anti-depressant prescription | WLGP | <https://conceptlibrary.saildatabank.com/concepts/C2917/detail/> |

### Table S4: Male to female ratios & sample sizes

| **Category** | **Group** | **Males** | **Females** | **Total** | **% total*** | **M:F ratio** |
| --- | --- | --- | --- | --- | --- | --- |
| **Everyone** | Everyone | 13110 | 3348 | 16458 | 100.0% | 3.9:1 |
| **Age at first recorded diagnosis** | <12 | 8798 | 1821 | 10619 | 64.5% | 4.8:1 |
|  | >=12 | 4312 | 1527 | 5839 | 35.5% | 2.8:1 |
|  | <18 | 12011 | 2775 | 14786 | 89.8% | 4.3:1 |
|  | >=18 | 1099 | 573 | 1672 | 10.2% | 1.9:1 |
| **Birth years** | 1989-1993 | 2393 | 680 | 3073 | 18.7% | 3.5:1 |
|  | 1994-1998 | 3277 | 877 | 4154 | 25.2% | 3.7:1 |
|  | 1999-2003 | 3035 | 807 | 3842 | 23.3% | 3.8:1 |
|  | 2004-2008 | 2956 | 669 | 3625 | 22.0% | 4.4:1 |
|  | 2009-2013 | 1449 | 315 | 1764 | 10.7% | 4.6:1 |
| **Year at first recorded diagnosis** | 2000-2003 | 1666 | 324 | 1990 | 12.1% | 5.1:1 |
|  | 2004-2007 | 2311 | 475 | 2786 | 16.9% | 4.9:1 |
|  | 2008-2011 | 2433 | 537 | 2970 | 18.0% | 4.5:1 |
|  | 2012-2015 | 2835 | 815 | 3650 | 22.2% | 3.5:1 |
|  | 2016-2019 | 3865 | 1197 | 5062 | 30.8% | 3.2:1 |
| **Ethnicity** | Ethnic majority (White) | 8761 | 2239 | 11000 | 97.1% | 3.9:1 |
|  | Ethnic minority** | 268 | 65 | 333 | 2.9% | 4.1:1 |
| **WIMD** | 1 (least deprived) | 1463 | 405 | 1868 | 11.6% | 3.6:1 |
|  | 2 | 1655 | 455 | 2110 | 13.1% | 3.6:1 |
|  | 3 | 2119 | 553 | 2672 | 16.6% | 3.8:1 |
|  | 4 | 3115 | 772 | 3887 | 24.1% | 4.0:1 |
|  | 5 (most deprived) | 4507 | 1096 | 5603 | 34.7% | 4.1:1 |
| **Social services involvement** | Looked after children (LAC) | 702 | 211 | 913 | 5.5% | 3.3:1 |
|  | Child protection register (CPR) | 511 | 142 | 653 | 4.0% | 3.6:1 |
|  | LAC and/or CPR | 1026 | 306 | 1332 | 8.1% | 3.4:1 |
| **Year at first recorded diagnosis - split by every year** | 2000 | 330 | 76 | 406 | 2.5% | 4.3:1 |
|  | 2001 | 356 | 57 | 413 | 2.5% | 6.2:1 |
|  | 2002 | 441 | 85 | 526 | 3.2% | 5.2:1 |
|  | 2003 | 539 | 106 | 645 | 3.9% | 5.1:1 |
|  | 2004 | 552 | 104 | 656 | 4.0% | 5.3:1 |
|  | 2005 | 565 | 127 | 692 | 4.2% | 4.4:1 |
|  | 2006 | 594 | 126 | 720 | 4.4% | 4.7:1 |
|  | 2007 | 600 | 118 | 718 | 4.4% | 5.1:1 |
|  | 2008 | 575 | 111 | 686 | 4.2% | 5.2:1 |
|  | 2009 | 579 | 116 | 695 | 4.2% | 5.0:1 |
|  | 2010 | 585 | 146 | 731 | 4.4% | 4.0:1 |
|  | 2011 | 694 | 164 | 858 | 5.2% | 4.2:1 |
|  | 2012 | 600 | 154 | 754 | 4.6% | 3.9:1 |
|  | 2013 | 720 | 192 | 912 | 5.5% | 3.8:1 |
|  | 2014 | 742 | 224 | 966 | 5.9% | 3.3:1 |
|  | 2015 | 773 | 245 | 1018 | 6.2% | 3.2:1 |
|  | 2016 | 915 | 274 | 1189 | 7.2% | 3.3:1 |
|  | 2017 | 1015 | 304 | 1319 | 8.0% | 3.3:1 |
|  | 2018 | 975 | 298 | 1273 | 7.7% | 3.3:1 |
|  | 2019 | 960 | 321 | 1281 | 7.8% | 3.0:1 |

* Percentages shown reflect the proportion within each category in individuals with ADHD with available data for that variable. ** Includes ONS-defined groups: Asian/Asian British, Black/African/Caribbean/Black British, mixed/multiple ethnic groups, and other ethnic minority group.

### Figure S1: Male to female ratio split by year at first recorded diagnosis

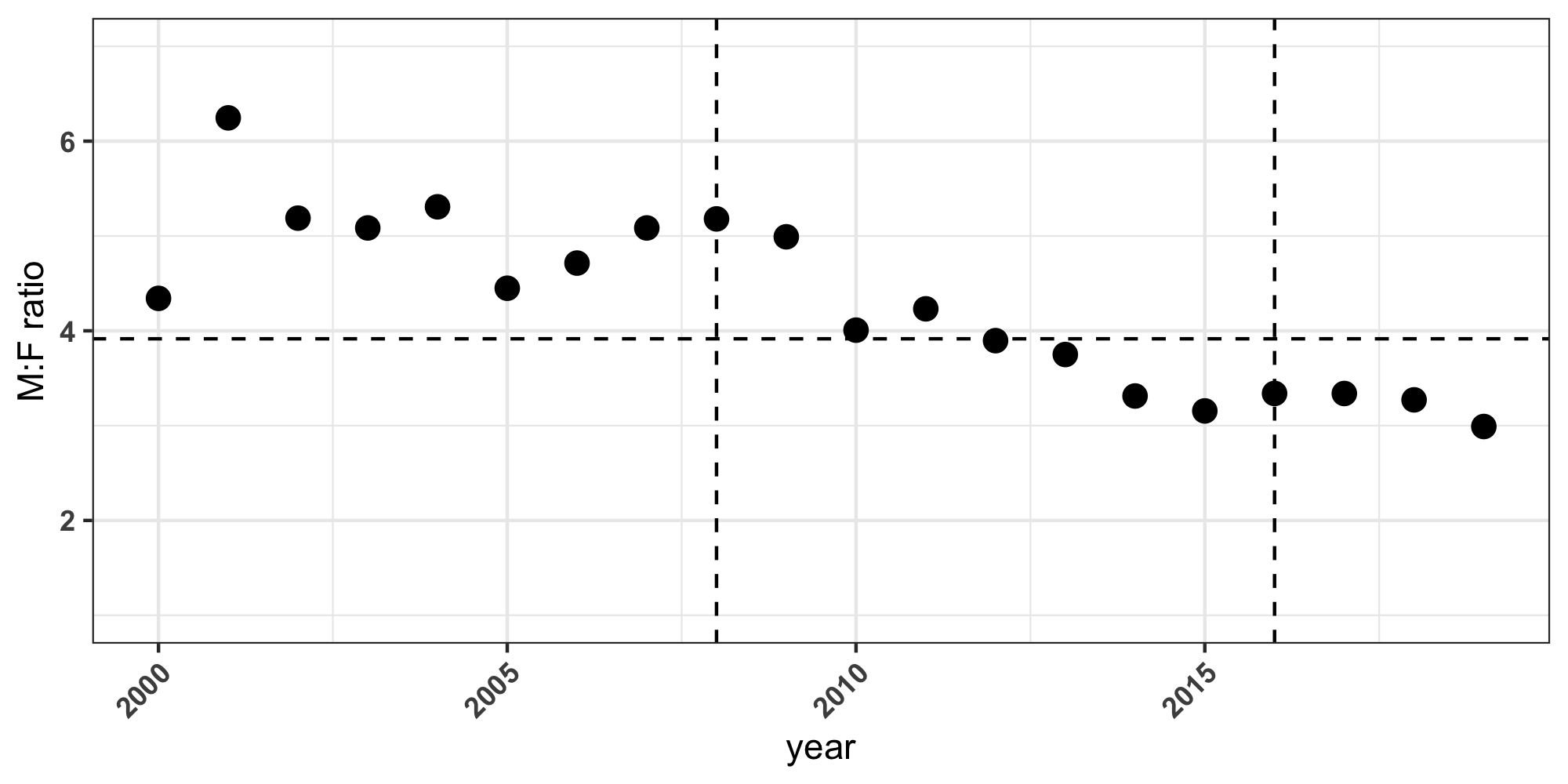

Dashed vertical lines indicate the years: 2008 (updated NICE guidelines) and 2016 (introduction of joint neurodevelopmental services in Wales)

### Table S5: Regression analyses testing for sex differences in ADHD diagnosis and clinical care

| **Variable (continuous)** | **N Males** | **N Females** | **Mean(SE) M** | **Mean(SE) F** | **OR (95% CI)*** | **p_FDR_** |
| --- | --- | --- | --- | --- | --- | --- |
| Age at first ADHD diagnosis | 13110 | 3348 | 10.90 (0.04) | 12.60 (0.10) | 1.08 (1.07-1.09) | 1.7E-60 |
| Age at first ADHD medication (any) | 8415 | 1972 | 11.20 (0.05) | 12.80 (0.12) | 1.09 (1.08-1.11) | 1.2E-42 |
| Age at first stimulant prescription | 8015 | 1873 | 11.10 (0.05) | 12.70 (0.12) | 1.10 (1.08-1.11) | 9.2E-44 |
| Age at first non-stimulant prescription | 1676 | 397 | 13.30 (0.12) | 14.50 (0.26) | 1.05 (1.02-1.08) | 9.8E-04 |
| Age at first anti-depressant medication prescription | 2478 | 1205 | 20.20 (0.09) | 19.80 (0.11) | 0.97 (0.95-0.99) | 2.3E-03 |
| Age at first ASD diagnosis | 1804 | 382 | 11.00 (0.12) | 12.20 (0.29) | 1.06 (1.03-1.09) | 8.2E-06 |
| Age at first LD diagnosis | 1097 | 346 | 11.40 (0.16) | 12.70 (0.32) | 1.04 (1.01-1.07) | 3.3E-03 |
| Age at first other mental health diagnosis | 4194 | 1242 | 14.20 (0.09) | 14.70 (0.14) | 1.01 (1.00-1.03) | 0.11 |
| Age at first anxiety diagnosis | 2251 | 1107 | 18.30 (0.11) | 18.50 (0.14) | 1.02 (1.00-1.04) | 0.071 |
| Age at first depression diagnosis | 2552 | 1255 | 19.80 (0.08) | 18.60 (0.10) | 0.90 (0.88-0.92) | 3.8E-20 |
| Age difference: ADHD diagnosis vs ADHD medication | 6685 | 1601 | 1.10 (0.02) | 1.09 (0.05) | 1.00 (0.97-1.02) | 0.73 |
| **Variable (binary)** | **N Males** | **N Females** | **N(%) M** | **N(%) F** | **OR (95% CI)** | **p_FDR_** |
| ADHD diagnosis group (<12 vs 12+) | 13110 | 3348 | 4312 (32.9) | 1527 (45.6) | 1.75 (1.60-1.91) | 1.1E-34 |
| ADHD diagnosis group (<18 vs 18+) | 13110 | 3348 | 1099 (8.4) | 573 (17.1) | 2.32 (2.04-2.63) | 4.5E-39 |
| Any ADHD medication | 13110 | 3348 | 8415 (64.2) | 1972 (58.9) | 0.79 (0.73-0.85) | 3.5E-09 |
| Any stimulants | 13110 | 3348 | 8015 (61.1) | 1873 (55.9) | 0.80 (0.74-0.86) | 1.4E-08 |
| Any non-stimulants | 13110 | 3348 | 1676 (12.8) | 397 (11.9) | 0.91 (0.80-1.02) | 0.11 |
| Both stimulants & non-stimulants | 13110 | 3348 | 1276 (9.7) | 298 (8.9) | 0.89 (0.78-1.02) | 0.11 |
| Any anti-depressant medication | 13110 | 3348 | 2478 (18.9) | 1205 (36.0) | 2.75 (2.50-3.03) | 1.6E-93 |
| Any other mental health diagnosis | 13110 | 3348 | 4194 (32.0) | 1242 (37.1) | 1.22 (1.12-1.32) | 5.7E-06 |
| Any anxiety diagnosis | 13110 | 3348 | 2251 (17.2) | 1107 (33.1) | 2.46 (2.24-2.69) | 1.9E-83 |
| Any depression diagnosis | 13110 | 3348 | 2552 (19.5) | 1255 (37.5) | 2.86 (2.60-3.15) | 1.7E-101 |
| Other mental health diagnosis first recorded prior to ADHD diagnosis | 4194 | 1242 | 1482 (35.3) | 583 (46.9) | 1.62 (1.42-1.84) | 4.0E-13 |
| Anxiety first recorded prior to ADHD diagnosis | 2251 | 1107 | 448 (19.9) | 330 (29.8) | 1.71 (1.44-2.02) | 6.4E-10 |
| Depression first recorded prior to ADHD diagnosis | 2552 | 1255 | 550 (21.6) | 446 (35.5) | 2.00 (1.72-2.33) | 1.4E-18 |
| Anti-depressant first recorded prior to ADHD diagnosis | 2478 | 1205 | 436 (17.6) | 344 (28.5) | 1.85 (1.57-2.19) | 1.4E-12 |
| Anti-depressant continuation after ADHD diagnosis (in those prescribed anti-depressants prior to ADHD diagnosis) | 436 | 344 | 207 (47.5) | 125 (36.3) | 0.63 (0.47-0.84) | 2.2E-03 |

* Males are the reference group. ASD: autism spectrum disorder; LD: learning difficulties; MH: mental health; FDR: false discovery rate correction.

### Table S6: Results of regression analyses comparing males and females with ADHD on clinical characteristics, in those with social services involvement (N=1,332)

| **Variable (continuous)** | **N M** | **N F** | **Mean(SE) M** | **Mean(SE) F** | **OR (95% CI)*** |
| --- | --- | --- | --- | --- | --- |
| Age at first ADHD diagnosis | 1026 | 306 | 9.60 (0.11) | 10.70 (0.22) | 1.10 (1.06-1.14) |
| Age at first ADHD medication (any) | 681 | 179 | 10.10 (0.12) | 10.80 (0.24) | 1.08 (1.02-1.14) |
| Age at first stimulant prescription | 648 | 165 | 10.00 (0.12) | 10.90 (0.25) | 1.09 (1.03-1.15) |
| Age at first non-stimulant prescription | 149 | 44 | 11.30 (0.26) | 12.10 (0.54) | 1.09 (0.98-1.22) |
| Age at first anti-depressant medication prescription | 141 | 77 | 17.60 (0.29) | 17.30 (0.37) | 0.93 (0.83-1.03) |
| Age at first ASD diagnosis | 153 | 33 | 9.46 (0.36) | 10.90 (0.63) | 1.10 (1.00-1.21) |
| Age at first LD diagnosis | 134 | 49 | 10.70 (0.41) | 11.60 (0.69) | 1.05 (0.97-1.14) |
| Age at first other mental health diagnosis | 413 | 154 | 12.50 (0.21) | 12.50 (0.32) | 0.98 (0.94-1.03) |
| Age at first anxiety diagnosis | 167 | 83 | 15.50 (0.34) | 16.50 (0.40) | 1.08 (0.99-1.19) |
| Age at first depression diagnosis | 144 | 93 | 17.60 (0.28) | 16.10 (0.28) | 0.83 (0.74-0.92) |
| Age difference: ADHD diagnosis vs ADHD medication | 559 | 151 | 1.20 (0.08) | 0.97 (0.11) | 0.92 (0.82-1.02) |
| **Variable (binary)** | **N M** | **N F** | **N(%) M** | **N(%) F** | **OR (95% CI)** |
| ADHD diagnosis group (<12 vs 12+) | 1026 | 306 | 237 (23.1) | 105 (34.3) | 1.80 (1.33-2.45) |
| ADHD diagnosis group (<18 vs 18+) | 1026 | 306 | 21 (2.1) | 17 (5.6) | 2.77 (1.39-5.49) |
| Any ADHD medication | 1026 | 306 | 681 (66.4) | 179 (58.5) | 0.70 (0.54-0.91) |
| Any stimulants | 1026 | 306 | 648 (63.2) | 165 (53.9) | 0.67 (0.52-0.87) |
| Any non-stimulants | 1026 | 306 | 149 (14.5) | 44 (14.4) | 0.95 (0.65-1.36) |
| Both stimulants & non-stimulants | 1026 | 306 | 116 (11.3) | 30 (9.8) | 0.82 (0.52-1.24) |
| Any anti-depressant medication | 1026 | 306 | 141 (13.7) | 77 (25.2) | 2.21 (1.56-3.12) |
| Any other mental health diagnosis | 1026 | 306 | 413 (40.3) | 154 (50.3) | 1.49 (1.13-1.95) |
| Any anxiety diagnosis | 1026 | 306 | 167 (16.3) | 83 (27.1) | 1.92 (1.39-2.64) |
| Any depression diagnosis | 1026 | 306 | 144 (14.0) | 93 (30.4) | 3.14 (2.22-4.45) |
| Other mental health diagnosis first recorded prior to ADHD diagnosis | 413 | 154 | 133 (32.2) | 72 (46.8) | 1.84 (1.26-2.69) |
| Anxiety first recorded prior to ADHD diagnosis | 167 | 83 | 31 (18.6) | 14 (16.9) | 0.90 (0.44-1.79) |
| Depression first recorded prior to ADHD diagnosis | 144 | 93 | 22 (15.3) | 26 (28.0) | 2.26 (1.13-4.60) |
| Anti-depressant first recorded prior to ADHD diagnosis | 141 | 77 | 14 (9.9) | 16 (20.8) | 2.19 (0.97-5.00) |
| Anti-depressant continuation after ADHD diagnosis | 14 | 16 | n<10** | n<10 | 0.49 (0.08-2.73) |

* Males are the reference group; ** Results with fewer than 10 individuals cannot be shown as per Safe Researcher guidelines. ASD: autism spectrum disorder; LD: learning difficulties; MH: mental health

### Table S7: Results of sensitivity analyses comparing males and females with ADHD on clinical characteristics, in the subgroup with ADHD after age 5 and complete information from age 5 onwards (N=12,301)

| **Variable (continuous)** | **N M** | **N F** | **Mean(SE) M** | **Mean(SE) F** | **OR (95% CI)*** |
| --- | --- | --- | --- | --- | --- |
| Age at first ADHD diagnosis | 9860 | 2441 | 10.00 (0.04) | 11.30 (0.09) | 1.09 (1.08-1.10) |
| Age at first ADHD medication (any) | 6353 | 1430 | 10.40 (0.04) | 11.50 (0.11) | 1.10 (1.08-1.12) |
| Age at first stimulant prescription | 6058 | 1362 | 10.30 (0.04) | 11.40 (0.11) | 1.10 (1.08-1.12) |
| Age at first non-stimulant prescription | 1249 | 277 | 11.80 (0.11) | 12.80 (0.24) | 1.08 (1.04-1.12) |
| Age at first anti-depressant medication prescription | 1250 | 644 | 18.20 (0.10) | 18.10 (0.12) | 0.98 (0.94-1.02) |
| Age at first ASD diagnosis | 1440 | 304 | 10.00 (0.12) | 10.90 (0.26) | 1.06 (1.03-1.10) |
| Age at first LD diagnosis | 825 | 260 | 10.20 (0.16) | 11.30 (0.32) | 1.05 (1.01-1.08) |
| Age at first other mental health diagnosis | 2644 | 800 | 12.50 (0.10) | 13.50 (0.16) | 1.03 (1.01-1.05) |
| Age at first anxiety diagnosis | 1287 | 674 | 15.80 (0.13) | 16.60 (0.15) | 1.05 (1.02-1.08) |
| Age at first depression diagnosis | 1246 | 681 | 17.90 (0.09) | 17.20 (0.11) | 0.91 (0.88-0.95) |
| Age difference: ADHD diagnosis vs ADHD medication | 5299 | 1190 | 1.02 (0.02) | 1.05 (0.05) | 1.01 (0.97-1.04) |
| **Variable (binary)** | **N M** | **N F** | **N(%) M** | **N(%) F** | **OR (95% CI)** |
| ADHD diagnosis group (<12 vs 12+) | 9860 | 2441 | 2518 (25.5) | 924 (37.9) | 1.80 (1.63-2.00) |
| ADHD diagnosis group (<18 vs 18+) | 9860 | 2441 | 414 (4.2) | 225 (9.2) | 2.27 (1.88-2.72) |
| Any ADHD medication | 9860 | 2441 | 6353 (64.4) | 1430 (58.6) | 0.76 (0.70-0.83) |
| Any stimulants | 9860 | 2441 | 6058 (61.4) | 1362 (55.8) | 0.78 (0.71-0.85) |
| Any non-stimulants | 9860 | 2441 | 1249 (12.7) | 277 (11.3) | 0.85 (0.74-0.98) |
| Both stimulants & non-stimulants | 9860 | 2441 | 954 (9.7) | 209 (8.6) | 0.84 (0.72-0.99) |
| Any anti-depressant medication | 9860 | 2441 | 1250 (12.7) | 644 (26.4) | 2.64 (2.34-2.98) |
| Any other mental health diagnosis | 9860 | 2441 | 2644 (26.8) | 800 (32.8) | 1.29 (1.17-1.42) |
| Any anxiety diagnosis | 9860 | 2441 | 1287 (13.1) | 674 (27.6) | 2.56 (2.29-2.86) |
| Any depression diagnosis | 9860 | 2441 | 1246 (12.6) | 681 (27.9) | 2.90 (2.57-3.27) |
| Other mental health diagnosis first recorded prior to ADHD diagnosis | 2644 | 800 | 953 (36.0) | 362 (45.2) | 1.49 (1.27-1.75) |
| Anxiety first recorded prior to ADHD diagnosis | 1287 | 674 | 245 (19.0) | 182 (27.0) | 1.59 (1.27-1.98) |
| Depression first recorded prior to ADHD diagnosis | 1246 | 681 | 238 (19.1) | 222 (32.6) | 2.03 (1.64-2.53) |
| Anti-depressant first recorded prior to ADHD diagnosis | 1250 | 644 | 174 (13.9) | 157 (24.4) | 1.93 (1.50-2.47) |
| Anti-depressant continuation after ADHD diagnosis | 174 | 157 | 92 (52.9) | 55 (35.0) | 0.48 (0.31-0.75) |

* Males are the reference group. ASD: autism spectrum disorder; LD: learning difficulties; MH: mental health

### Table S8: Results of sensitivity analyses comparing males and females with ADHD on clinical characteristics, in the subgroup with good coverage across the study period (N=9,816)

| **Variable (continuous)** | **N M** | **N F** | **Mean(SE) M** | **Mean(SE) F** | **OR (95% CI)*** |
| --- | --- | --- | --- | --- | --- |
| Age at first ADHD diagnosis | 7904 | 1912 | 9.32 (0.04) | 10.30 (0.09) | 1.08 (1.07-1.10) |
| Age at first ADHD medication (any) | 4929 | 1098 | 9.89 (0.04) | 10.90 (0.11) | 1.09 (1.07-1.12) |
| Age at first stimulant prescription | 4706 | 1044 | 9.87 (0.05) | 10.90 (0.11) | 1.09 (1.07-1.12) |
| Age at first non-stimulant prescription | 949 | 213 | 11.10 (0.11) | 12.10 (0.26) | 1.09 (1.04-1.14) |
| Age at first anti-depressant medication prescription | 833 | 414 | 17.60 (0.12) | 17.60 (0.14) | 0.98 (0.94-1.03) |
| Age at first ASD diagnosis | 1179 | 262 | 9.25 (0.12) | 10.30 (0.27) | 1.07 (1.04-1.11) |
| Age at first LD diagnosis | 684 | 203 | 9.49 (0.16) | 10.30 (0.35) | 1.03 (0.99-1.07) |
| Age at first other mental health diagnosis | 1972 | 587 | 11.60 (0.11) | 12.60 (0.18) | 1.03 (1.01-1.06) |
| Age at first anxiety diagnosis | 905 | 476 | 14.90 (0.15) | 15.80 (0.18) | 1.04 (1.00-1.08) |
| Age at first depression diagnosis | 794 | 447 | 17.30 (0.11) | 16.60 (0.13) | 0.90 (0.86-0.95) |
| Age difference: ADHD diagnosis vs ADHD medication | 4207 | 927 | 1.08 (0.03) | 1.08 (0.06) | 0.99 (0.95-1.03) |
| **Variable (binary)** | **N M** | **N F** | **N(%) M** | **N(%) F** | **OR (95% CI)** |
| ADHD diagnosis group (<12 vs 12+) | 7904 | 1912 | 1571 (19.9) | 593 (31.0) | 1.81 (1.60-2.04) |
| ADHD diagnosis group (<18 vs 18+) | 7904 | 1912 | 169 (2.1) | 99 (5.2) | 2.32 (1.78-3.01) |
| Any ADHD medication | 7904 | 1912 | 4929 (62.4) | 1098 (57.4) | 0.79 (0.72-0.88) |
| Any stimulants | 7904 | 1912 | 4706 (59.5) | 1044 (54.6) | 0.80 (0.72-0.88) |
| Any non-stimulants | 7904 | 1912 | 949 (12.0) | 213 (11.1) | 0.88 (0.75-1.03) |
| Both stimulants & non-stimulants | 7904 | 1912 | 726 (9.2) | 159 (8.3) | 0.86 (0.72-1.03) |
| Any anti-depressant medication | 7904 | 1912 | 833 (10.5) | 414 (21.7) | 2.46 (2.13-2.84) |
| Any other mental health diagnosis | 7904 | 1912 | 1972 (24.9) | 587 (30.7) | 1.29 (1.15-1.44) |
| Any anxiety diagnosis | 7904 | 1912 | 905 (11.4) | 476 (24.9) | 2.56 (2.25-2.92) |
| Any depression diagnosis | 7904 | 1912 | 794 (10.0) | 447 (23.4) | 2.92 (2.53-3.36) |
| Other mental health diagnosis first recorded prior to ADHD diagnosis | 1972 | 587 | 731 (37.1) | 264 (45.0) | 1.41 (1.17-1.70) |
| Anxiety first recorded prior to ADHD diagnosis | 905 | 476 | 175 (19.3) | 130 (27.3) | 1.63 (1.25-2.12) |
| Depression first recorded prior to ADHD diagnosis | 794 | 447 | 128 (16.1) | 131 (29.3) | 2.18 (1.65-2.88) |
| Anti-depressant first recorded prior to ADHD diagnosis | 833 | 414 | 89 (10.7) | 79 (19.1) | 1.96 (1.41-2.73) |
| Anti-depressant continuation after ADHD diagnosis | 89 | 79 | 50 (56.2) | 23 (29.1) | 0.30 (0.15-0.57) |

* Males are the reference group. ASD: autism spectrum disorder; LD: learning difficulties; MH: mental health
